# Artificial Intelligence Generated Computed Tomography Segmentation of the Thoracoabdominal Aorta

**DOI:** 10.1101/2025.01.13.25320502

**Authors:** Devina Chatterjee, Nicholas T. Obey, Benjamin Shou, Sangmita Singh, Adrian R. Acuna Higaki, Adham Ahmed, Ely Erez, Michela Cupo, Nathaniel Price, Irbaz Hameed, Eric B. Schneider, Prashanth Vallabhajosyula, Chin Siang Ong

**Author notes:** **Corresponding authors:** Chin Siang Ong, MBBS, PhD, Assistant Professor of Surgery, Division of Surgical Outcomes, Surgery Center for Health Services and Outcomes Research, Department of Surgery, Yale School of Medicine, 100 Church Street South, New Haven, CT 06519, Email Address, Prashanth Vallabhajosyula, MD, MS, Director, Yale Aortic Institute, Director, Yale Pulmonary Thromboendarterectomy Program, Division of Cardiac Surgery, Yale School of Medicine, 330 Cedar Street, BB 204, New Haven, CT 06510. These authors contributed equally to this work (Prashanth Vallabhajosyula, Chin Siang Ong).

## Abstract

**Objectives:** The rising global burden of cardiovascular diseases (CV) highlights the critical need for efficiency in disease diagnosis and management. An important area for such improvement is utilization of artificial intelligence (AI) for streamlining time and resources in CV imaging workflow. We evaluate the performance of artificial intelligence (AI) segmentation for aortic segmentation on clinical computed tomography angiography (CTA) images and compare accuracy to manual methods. Such automation would markedly improve efficiency and accuracy of aortic surveillance.

**Methods:** This retrospective study included 27 scans from 20 patients who underwent thoracic endovascular aortic repair (TEVAR) between January 2020 and March 2022. An open-source AI model was applied to segment the aorta, and its performance was assessed by comparing AI-generated segmentations with manual segmentations using Dice similarity coefficients, volumetric analysis, and aortic dimensions. Centerline reconstructed images of thoracoabdominal aorta were processed to extract radiomic features, including maximum diameter and cross-sectional area, for analysis.

**Results:** The AI tool achieved a median Dice coefficient of 0.96 (0.02), indicating a high degree of concordance with manual segmentation. Multiplanar reconstruction was performed to visualize the aorta and extract measurements along its length using the automated centerline, and radiomic features, including maximum diameter and cross-sectional area, were subsequently extracted for analysis.

**Conclusions:** AI segmentation demonstrates strong potential for improving efficiency and consistency in thoracoabdominal aortic segmentation, achieving high accuracy compared to manual methods. These findings highlight the feasibility of AI integration into clinical practice for diagnosis and surveillance of aortopathies, warranting further validation on larger datasets to enable clinical translation.

**Graphical Abstract:** 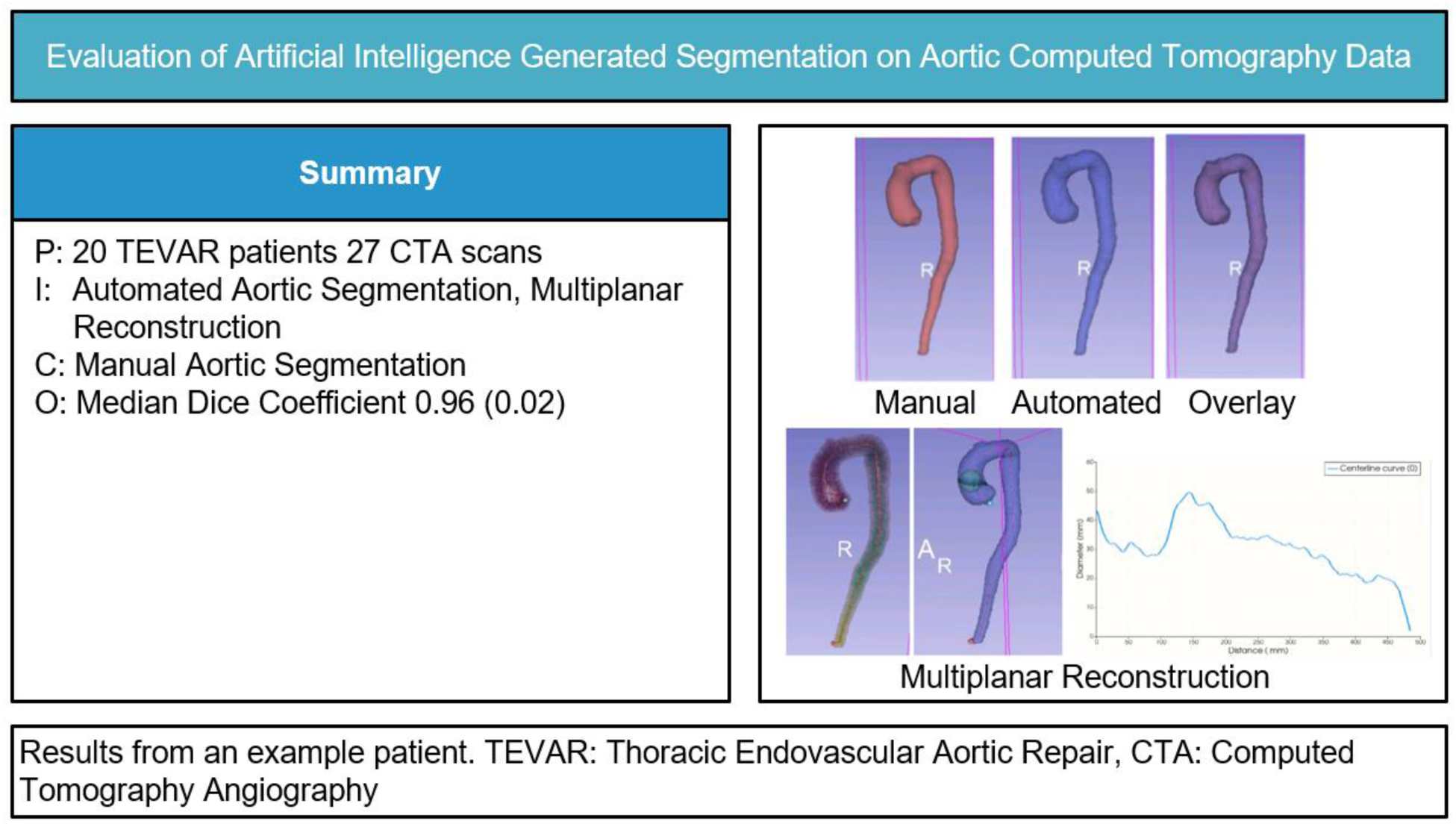

## Introduction

Imaging modalities play a critical role in diagnosis, surveillance, and clinical decision-making of cardiovascular (CV) pathologies, which affect a large portion of the population. Therefore, significant health system and health care provider (HCP) time and resources are dedicated to ever increasing CV imaging demands. Typically, clinicians manually outline regions of interest on images—is time-consuming, labor-intensive, and prone to variability [1–2]. At least 6 in 10 U.S. adults (61%), more than 184 million people, are expected to have some form of cardiovascular disease (CVD) within the next 30 years, contributing to a staggering $1.8 trillion in direct and indirect costs [3]. Given the prevalence of CVD and the necessity of regular imaging-based risk assessments, manual segmentation is becoming increasingly impractical for large-scale studies and clinical settings. It is time-consuming, prone to interpreter discordance and variation, increasing demands for 24 hour imaging review may introduce HCP fatigue and propensity for errors. These points underscore the critical need for reliable, automated systems to supplement and complement the current demands faced by CV radiologists and HCPs to achieve wide-scale application across our health system [4–6].

Automated segmentation offers a promising alternative, addressing the drawbacks of manual methods while maintaining, or even exceeding, their benefits, such as predictive value, risk stratification, and standardization [2]. Moreover, automated segmentation reduces observer bias, is more scalable, and requires less time [1]. For AI technologies to be implemented in clinical practice, they must be validated using real-world clinical data, as we aimed to do. One such area in CVD where clinical decision making is heavily dependent on imaging modalities is the diagnosis and management of aortopathies. Patients with aortic diseases also need life-long surveillance with serial imaging to monitor for disease progression and integrity of aortic interventions. Therefore, in this study, we assessed the potential utility of AI segmentation of thoracoabdominal aorta CT angiography in patients undergoing complex hybrid aortic arch and descending thoracoabdominal repairs to enable automated accurate dimensional analysis compared to traditional, manual segmentation CT angiography analysis.

This study investigates the performance of AI segmentation tools applied to uncurated clinical data. The primary objectives were: (1) to assess the fidelity of AI-generated aortic segmentations against manual benchmarks, and (2) to identify the practical challenges and opportunities for clinical adoption of AI segmentation tools.

## Materials and Methods

### Patient Cohort

Patients who underwent hybrid arch repair with zones 0 to 5 thoracic endovascular aortic repair (TEVAR) at Yale School of Medicine from January 2020 to March 2022 were included [8], contributing 27 scans.

### AI Segmentation

CTA images were sourced in DICOM format and converted to NIfTI using the dicom2nifti Python package and segmented using TotalSegmentator V2, an nnU-Net based model designed for the segmentation of distinct anatomical structures in CT images, including the aorta. This model had shown a high Dice similarity coefficient, measuring the similarity between manual and automated measurements on a scale of 0 (no similarity) to 1 (identical sets), of 0.943 on diverse clinical data sets, indicating superior segmentation accuracy. The model has been trained and validated on a dataset comprising 1,204 CT examinations, which included images with various abnormalities and from different clinical settings, thereby ensuring robustness and reliability [8].

### Morphology Analysis

Segmentation measurements were performed using modules from the VMTK plug-in for 3DSlicer, an open-source platform for medical image informatics, image processing, and three-dimensional visualization [9]. The Extract Centerline module was first used to compute the centerline between endpoints placed at the aortic root and bifurcation. Multiplanar reconstruction (MPR) was then performed to visualize the aorta in different planes. The maximum inscribed sphere method was applied to measure aortic diameters accurately across various sections, and the Cross-Section Analysis module was used to generate the diameter and cross-section area of the segmentation at each point along the centerline. The accuracy of AI-driven aortic segmentation was evaluated using the Dice similarity coefficient, volumetric analysis, and the assessment of aortic dimensions. Radiomic features of interest were compared against manual segmentation results for two example patients to assess the capability of the automated method to achieve high fidelity in anatomical replication. Dice scores were calculated using the Segment Comparison module from the SlicerRT extension [10]. Statistics were generated using the Segmentation Statistics module, presented as median and interquartile range (IQR). Mann-Whitney U tests were used for comparisons, with p values less than 0.05 considered significant.

## Results

### Patient Cohort Characteristics

A total of 20 patients, predominantly males (70%), with a mean age of 58.5 ± 11.8 years, who underwent hybrid arch repair with TEVAR were included in this study, with a total of 27 scans (Table 1). The majority of patients presented with type A aortic dissections (70%), with common comorbidities such as hypertension (90%), dyslipidemia (50%), and diabetes (30%). This patient population was chosen for the analysis because it represents one of the most complex groups of patients to surveillance from an imaging standpoint, and clinical decision making is heavily dependent on findings on CTA imaging.

**Table 1.** Patient Demographics.

| Characteristic | Count (%) * |
| --- | --- |
| Age, mean (SD), years | 58.0 (11.8) |
| Male | 14 (70%) |
| Black | 4 (20%) |
| Hispanic | 1 (5%) |
| White | 15 (75%) |
| Height, mean (SD), cm | 174.3 (10.5) |
| Weight, mean (SD), kg | 96.0 (14.8) |
| Body mass index, mean (SD) | 31.9 (6.0) |
| Diabetes | 6 (30%) |
| Dyslipidemia | 10 (50%) |
| Hypertension | 18 (90%) |
| Smoking | 11 (55%) |
| Chronic obstructive pulmonary disease | 2 (12%) |
| Prior coronary artery bypass grafting surgery | 2 (10%) |
| Prior aortic surgery | 10 (50%) |
| Presenting etiology |  |
| Aneurysm | 1 (5%) |
| Type A aortic dissection | 14 (70%) |
| Type A/type B aortic dissection | 4 (20%) |
| Type B aortic dissection | 1 (5%) |
| Acuity |  |
| Acute | 10 (50%) |
| Chronic | 9 (45%) |
| Subacute | 1 (5%) |
| Status |  |
| Elective | 8 (40%) |
| Emergent | 4 (20%) |
| Urgent | 8 (40%) |

\* Unless otherwise specified

AI segmentations achieved a median Dice coefficient of 0.96 (0.02) (Table 2). Radiomic features obtained by AI segmentation were not significantly different from those obtained by manual segmentation (Table 3). This close agreement indicates the robustness of AI segmentation across varied patient anatomies.

**Table 2:** Comparison of AI and Manual Aortic Segmentation (Metrics)

| Metric | Median (IQR) |
| --- | --- |
| Dice Similarity Coefficient | 0.96 (0.02) |
| Percentage of True Positives | 1.14 (7.41) |
| Percentage of True Negatives | 98.77 (8.06) |
| Percentage of False Positives | 0.04 (0.07) |
| Percentage of False Negatives | 0.06 (0.32) |

**Table 3:**
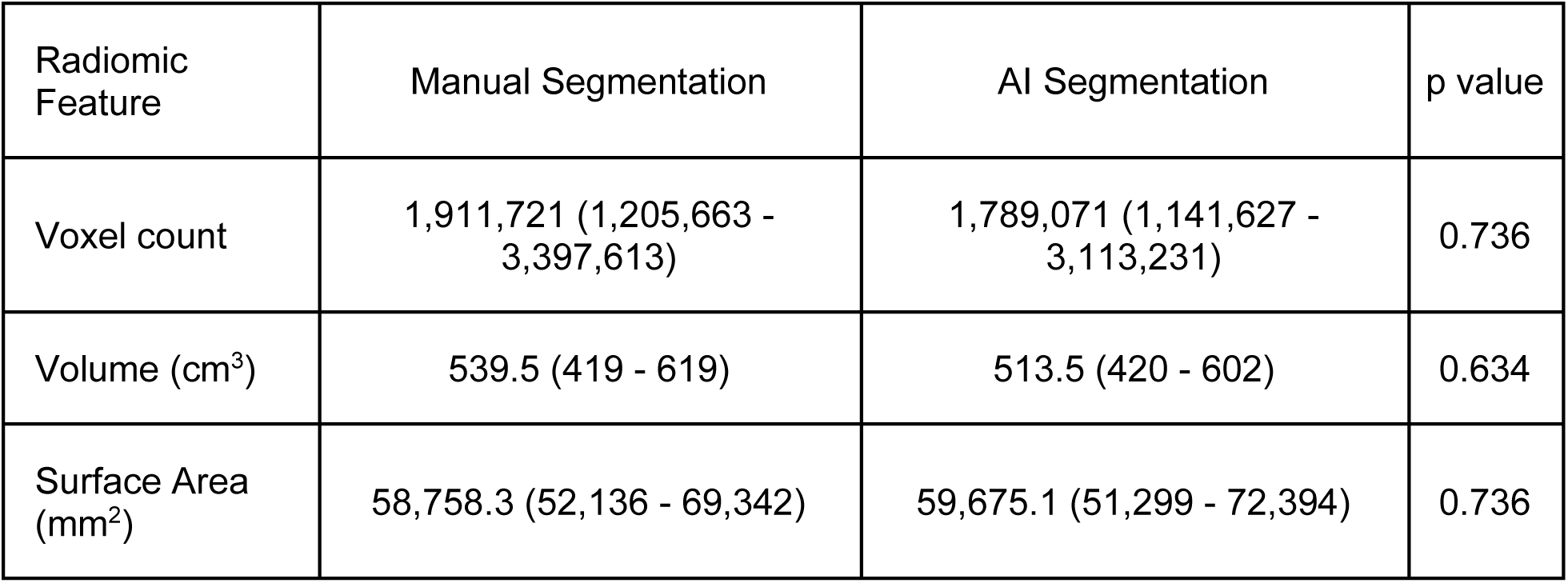
Comparison of AI and Manual Aortic Segmentation (Radiomic Features, Median (IQR))

### Quantitative Assessment of Aortic Segmentation Accuracy

The Dice similarity coefficient of Patient 1 was 0.92, indicating a high degree of concordance between the manual and AI-generated segmentations. The largest diameter along the aorta was recorded at 49.5 mm, with a total aortic distance from the root of 145.5 mm. AI segmentation produced a total volume of 481.8 cm³, which was 78 cm³ less than the volume determined manually. The voxel count for AI segmentation was 3,359,224, showing a reduction of 544,242 compared to the manual count.

The segmentation of Patient 2’s aorta showed an improved Dice similarity score of 0.97. The largest diameter was measured at 43.4 mm, and the total distance from the root was 68.5 mm. The AI model reported a volume of 450.2 cm³, 7 cm³ higher than the manual segmentation. The voxel count, representing the number of 3D volume units used to define the aorta, was 2,533,435—39,195 more than manual segmentation—indicating greater precision. The overlays presented in Figure 1 visually substantiate the segmentation accuracy, with AI-generated segmentations closely aligning with those manually delineated.

**Figure 1:**
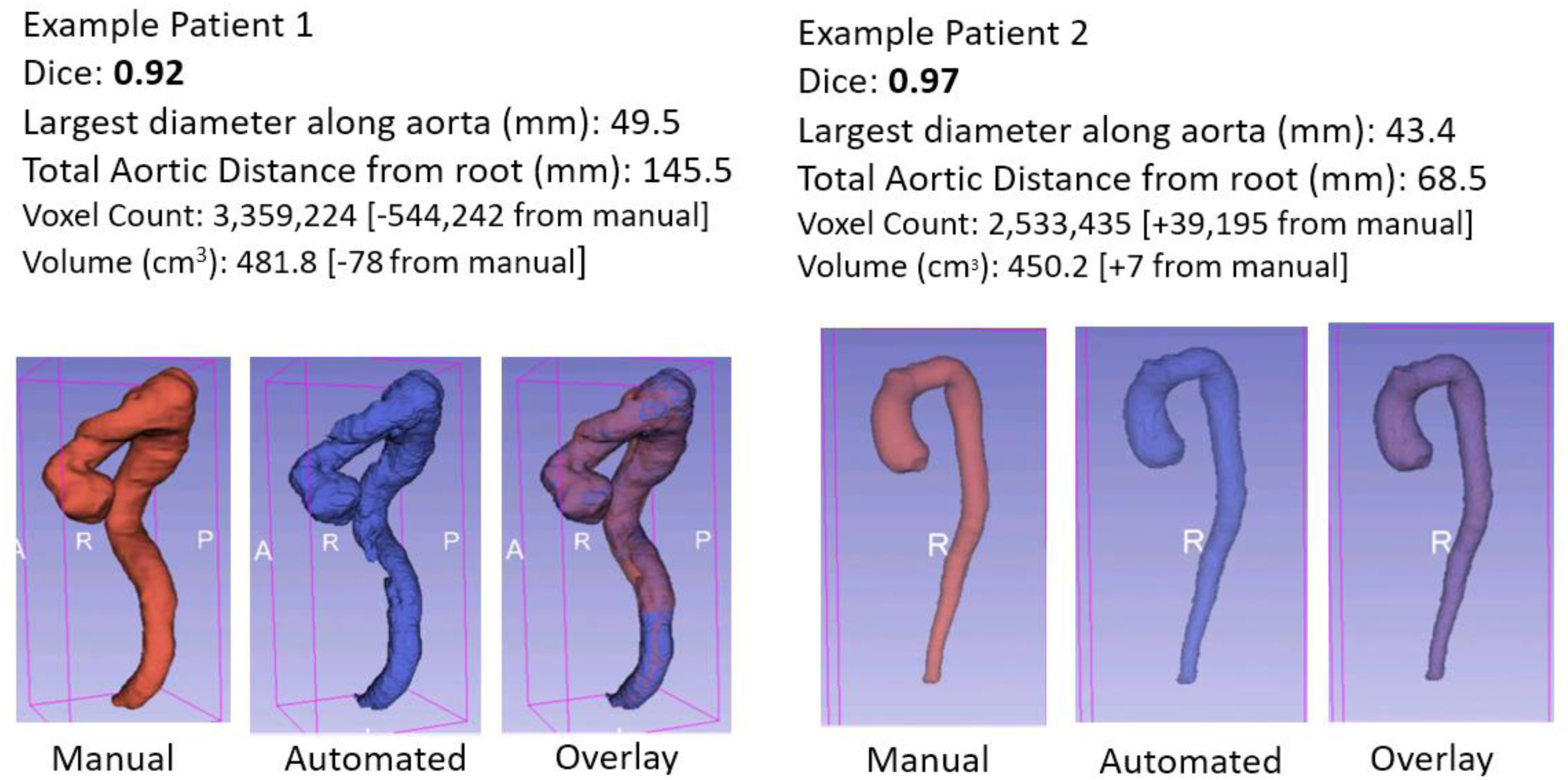
This figure illustrates the results of aortic segmentations from two example patients using manual and automated methods. The left panel shows results for Example Patient 1 and the right panel for Example Patient 2. Each panel is divided into three sub-panels displaying the manually segmented model, the automated segmented model, and an overlay of both, indicating comparative spatial alignment and segmentation consistency.

### Radiomic Feature Extraction from Segmented Models

Multiplanar reconstruction of the aorta was performed on Example Patient 2 using automated segmentation techniques (Figure 2). The centerline is used as a guide for extracting the aortic diameter at various points along its length, demonstrated in the right panel. The automated centerline extraction, performed using the maximum inscribed sphere method, is crucial for accurate measurement of the aortic diameter. This method provides an advantage over the manual method, which relies on the three orthogonal planes (sagittal, coronal, and axial) and cannot measure the aortic diameter based on the maximum inscribed sphere method without reconstruction. A plot of the unprocessed aortic diameter extracted from cross-sectional analysis along the centerline path.

**Figure 2:**
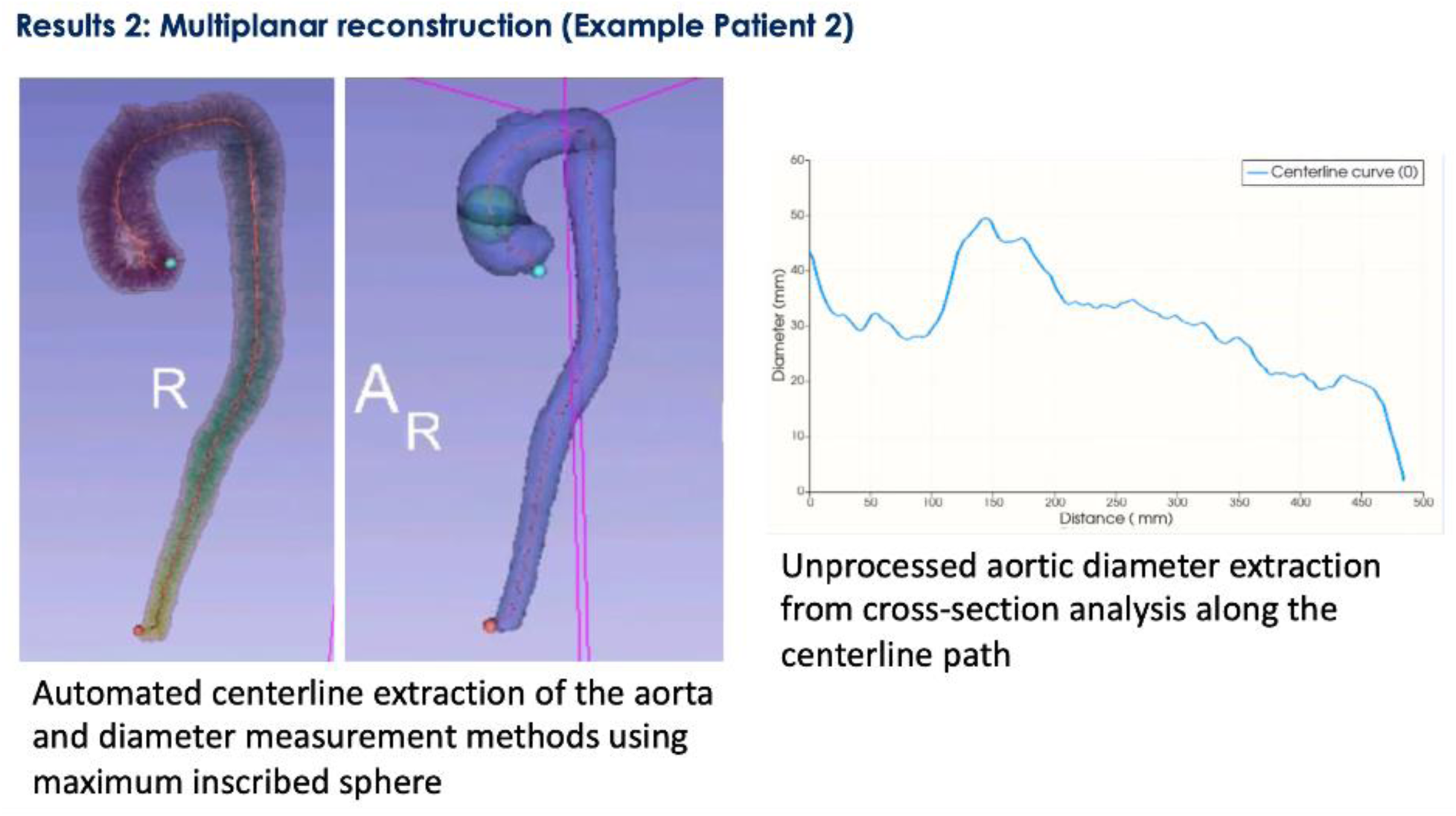
Multiplanar Reconstruction and Diameter Analysis for Example Patient 2. This figure demonstrates the advanced capabilities of automated segmentation and multiplanar reconstruction for detailed aortic analysis. Left Panel: Displays a 3D reconstruction of the aorta with automated centerline extraction, using the maximum inscribed sphere method to measure aortic diameters accurately across various sections. Right Panel: Presents a graph of unprocessed aortic diameters, extracted from cross-sectional analysis along the centerline. This graph delineates the changes in diameter along the aorta’s length, highlighting potential areas of pathological dilation or constriction crucial for surgical assessment and planning.

## Discussion

This study demonstrates the potential of AI to enhance the efficiency and accuracy of aortic segmentation. The high Dice coefficients observed indicate strong concordance between AI and manual segmentations. The differences in radiomic features obtained by AI and manual segmentation, e.g. volume and voxel counts, were not statistically significant. These results underscore AI’s ability to streamline the segmentation process while maintaining expert-level precision, significantly reducing the time and variability associated with manual methods, and still achieving high spatial overlap, as indicated by the Dice coefficients.

### Benefits of AI segmentation

AI segmentation offers several distinct advantages over manual methods. It eliminates inter-rater variability, providing consistent and reproducible results. Furthermore, it allows for high-throughput processing, enabling rapid analysis of large datasets, which is particularly useful in clinical settings with heavy workloads. AI’s scalability also ensures applicability across diverse patient populations and clinical conditions. Our aforedescribed approach, validated in patients with both normal aortas and thoracoabdominal pathology, is uniquely positioned to streamline analyses of pathological aortas and patient diagnoses. Automated segmentation has been rigorously validated in pathological aortic conditions, not just healthy tissue, including cases of aneurysms, dissections, and atherosclerotic disease. This ensures its reliability and accuracy in capturing clinically relevant variations in diameter, including peaks and troughs indicating dilations and constrictions, which are critical for surgical planning and risk assessment. These validations strengthen the clinical utility of the model, particularly in high-risk populations (Figure 2).

### Radiomics Advancements

Radiomics, supported by AI segmentation, offers detailed quantitative imaging features that extend beyond traditional measurements. For example, the ability to compute the maximum aortic diameter using a 3D centerline model enhances accuracy compared to manual measurements taken on sagittal, coronal, or transverse planes, which may misrepresent true cross-sectional diameter. Additionally, features such as shape descriptors, intensity scores, and volumes can provide valuable insights into aortic morphology and pathology. The high-throughput extraction of these features facilitates their integration into machine learning models, paving the way for advanced predictive analytics.

### Limitations

This study is a proof of concept, limited by a small sample size (n=27), which restricts the generalizability of the findings. Furthermore, integrating AI into clinical workflows poses significant challenges, including approval from regulatory bodies, clinician training, and compatibility with electronic health record systems. Additional validation and testing are needed to ensure seamless operation without disrupting existing workflow. Addressing these barriers is crucial for achieving widespread clinical adoption and ensuring effective implementation of AI tools.

## Conclusion

Pretrained, “out-of-the-box” AI tools exhibit promising accuracy in aortic segmentation from heterogeneous clinical data. These tools can potentially reduce the time and labor costs associated with manual segmentation, facilitating more detailed and rapid preoperative planning. Further studies are required to validate these findings across broader populations and to address the workflow and regulatory challenges identified.

## Author Contributions

Conceptualization: DC, BS, CSO, PV.

Data Curation: NTO, AAH, AA, IH, EE, MC, NP.

Formal Analysis: DC, NTO. Investigation: DC, NTO.

Methodology: DC, BS, NTO, CSO.

Project Administration: CSO.

Resources: NP, EBS, CSO.

Software: DC, NTO.

Supervision: EBS, IH, PV, CSO.

Visualization: DC, NTO, SS.

Writing – Original Draft: DC, BS, SS.

Writing – Review & Editing: EBS, IH, PV, CSO.

## Ethical Statement

This study was determined to be exempt from review by Yale University’s Institutional Review Board (IRB) under protocol number 2000032492, with Waiver of Consent/Waiver of HIPAA Authorization included in the study protocol due to minimal risk to participants and impracticality.

## Data Availability Statement

The data underlying this article cannot be shared publicly due to the privacy of individuals that participated in the study. Study data are available upon reasonable request to the corresponding author, in accordance with institutional policies and any applicable data sharing or data use agreements.

## References

[1] C. Ozturk, D. H. Pak, L. Rosalia, D. Goswami, M. E. Robakowski, R. McKay, et al., “AI-powered multimodal modeling of personalized hemodynamics in aortic stenosis,” ArXiv Prepr. ArXiv240700535, 2024.

[2] K. Liu, D. Zhao, L. Feng, Z. Zhang, P. Qiu, X. Wu, et al., “Unraveling phenotypic heterogeneity in stanford type B aortic dissection patients through machine learning clustering analysis of cardiovascular CT imaging,” Hellenic J. Cardiol., 2024.

[3] K. J. Maddox, M. S. Elkind, H. J. Aparicio, Y. Commodore-Mensah, S. D. de Ferranti, W. N. Dowd, et al., “Forecasting the burden of cardiovascular disease and stroke in the United States through 2050—prevalence of risk factors and disease: a presidential advisory from the American Heart Association,” Circulation, vol. 149, pp. e00–e00, 2024.

[4] C. Caradu, A.-L. Pouncey, E. Lakhlifi, C. Brunet, X. Bérard, and E. Ducasse, “Fully automatic volume segmentation using deep learning approaches to assess aneurysmal sac evolution after infrarenal endovascular aortic repair,” J. Vasc. Surg., vol. 76, no. 3, pp. 620–630.e3, 2022, doi: 10.1016/j.jvs.2022.03.891.

[5] C. Chen, C. Qin, H. Qiu, G. Tarroni, J. Duan, W. Bai, et al., “Deep learning for cardiac image segmentation: a review,” Front. Cardiovasc. Med., vol. 7, p. 25, 2020.

[6] H. Kim, J. Jung, J. Kim, B. Cho, J. Kwak, J. Y. Jang, et al., “Abdominal multi-organ auto-segmentation using 3D-patch-based deep convolutional neural network,” Sci. Rep., vol. 10, no. 1, p. 6204, 2020.

[7] I. Hameed, A. Ahmed, S. Pupovac, N. Nassiri, R. Assi, and P. Vallabhajosyula, “Aortic remodeling following hybrid arch repair with zone 0 to 5 thoracic endovascular aortic repairs for complex arch and descending thoracic aortic pathologies,” JTCVS Open, vol. 17, pp. 23– 36, Feb. 2024, doi: 10.1016/j.xjon.2023.12.004.

[8] J. Wasserthal, H.-C. Breit, M. T. Meyer, M. Pradella, D. Hinck, A. W. Sauter, et al., “TotalSegmentator: Robust Segmentation of 104 Anatomic Structures in CT Images,” *Radiol*. Artif. Intell., vol. 5, no. 5, 2023, doi: 10.1148/ryai.230024.

[9] L. Antiga, M. Piccinelli, L. Botti, B. Ene-Iordache, A. Remuzzi, and D. A. Steinman, “An image-based modeling framework for patient-specific computational hemodynamics,” Med. Biol. Eng. Comput., vol. 46, pp. 1097–1112, 2008.

[10] C. Pinter, A. Lasso, A. Wang, D. Jaffray, and G. Fichtinger, “SlicerRT: radiation therapy research toolkit for 3D Slicer,” Med. Phys., vol. 39, no. 10, pp. 6332–6338, 2012.

